# Evolution and Viral Properties of the SARS-CoV-2 BA.3.2 Subvariant

**DOI:** 10.1101/2025.10.28.25338622

**Authors:** Zesuliwe Jule, Cornelius Römer, Taskeen Hossen, Victoria Sviridchik, Kajal Reedoy, Yashica Ganga, Siphokazi Silangwe, Alex Norman, Farina Karim, Dikeledi Kekana, Boitshoko Mahlangu, Anele Mnguni, Ayanda Nzimande, Nadine Stock, Mallory Bernstein, Bernadett I. Gosnell, Mahomed-Yunus S. Moosa, Nicole Wolter, Khadija Khan, Richard A. Neher, Alex Sigal

## Abstract

The SARS-CoV-2 Omicron subvariant BA.3.2 descends from BA.3. It emerged 2 years after BA.3 ceased to circulate and differs by 39 spike mutations from BA.3. Similar to BA.2.86, which circulated at low levels before giving rise to JN.1, BA.3.2 shows a low but persistent circulation globally. Here, we characterize the phylogenetic origin, infection in cell culture, and neutralization of BA.3.2 using live virus and blood plasma samples collected in South Africa at different stages of the Covid-19 pandemic. Like the Omicron BA.2.86 subvariant, we find that BA.3.2 likely emerged in Southern Africa. We also find that in almost all BA.3.2 sequences to date, an 870 base pair deletion removes ORF7 and ORF8. In cell culture, BA.3.2 has lower cytotoxicity measured as plaque area compared to ancestral SARS-CoV-2, the Delta variant, and Omicron BA.1, but similar to the co-circulating LP.8.1 Omicron subvariant with whom it also shares similar replication in H1299-ACE2 cells. BA.3.2 and LP.8.1 exhibit complete escape from neutralization from pre-Omicron collected plasma samples, have low levels of neutralization by plasma collected in 2024, and higher neutralization by plasma collected in 2025, with BA.3.2 having moderately higher escape relative to LP.8.1. The emergence of long branch subvariants like BA.3.2 without intermediates likely indicates that unmonitored persistent infections continue to drive large evolutionary shifts in this virus.

## Introduction

The ongoing diversification of the SARS-CoV-2 Omicron lineage has produced multiple sub-lineages with distinct biological and immunological features and this process seems to continue in the face of rising levels of immunity to SARS-CoV-2^1^. BA.3.2 is a recently emerged Omicron subvariant first detected in South Africa^2^. BA.3.2 is a descendent of BA.3, an Omicron subvariant with limited circulation which first emerged in Southern Africa in late 2021, concurrently with BA.1 and BA.2^3^. Strikingly, the BA.3 variant last circulated in 2022^4^. It showed relatively limited spread, mostly in the South African region^5^. BA.3.2 contains 51 mutations relative to the BA.3 subvariant. So far, it caused relatively few confirmed infections from November 2024 when it was first detected, but these are widely distributed, including in South Africa, Australia, the Netherlands, and the USA^6^. Recent wastewater surveillance in Australia shows it is still present^7^.

While their origin cannot be conclusively proven, saltations such as BA.2.86 and BA.3.2, which have large numbers of mutations and no known circulating intermediates, are thought to evolve in persistent infections in immunocompromised people^1^. Given that SARS-CoV-2 and other coronaviruses have proofreading activity, the frequency of mutations is low relative to other RNA viruses^8^, and persistent infections are a mechanism to accumulate mutations which are otherwise lost at the transmission bottleneck in short, acute infections^1^. The evolutionary dynamics of BA.3.2 are similar to BA.2.86 which emerged in 2024^9^. The number of mutations is roughly comparable. Similarly to BA.3.2, BA.2.86 did not initially lead to many infections, but addition of the L455S substitution in spike led to the JN.1 sub-variant, which subsequently dominated SARS-CoV-2 infections globally^10^. Examining the origin of BA.2.86, we found that the closest ancestor was BA.2 circulating in Southern Africa in 2021-2022 and the closest BA.2.86 to the root of the BA.2.86 branch was also found in Southern Africa^9^. This makes it likely that BA.2.86 evolved in the Southern African region. Other variants and subvariants thought evolve in Southern Africa are Beta, BA.1, BA.2, and BA.3, making the region a hotspot for long-branch variants^1^. This concentration may be explained by the high prevalence of immunocompromise from advanced HIV disease in some regions of Southern Africa^1^.

There have been two previous studies which used the spike protein from BA.3.2 in a pseudotyped virus system to examine BA.3.2 cell entry and escape from neutralizing antibodies^2,11^. The results showed that BA.3.2 spike had higher escape from neutralizing antibodies in sera from participants with various histories of exposure to SARS-CoV-2 from vaccination when compared with the co-circulating LP.8.1^2,11^. However, BA.3.2 spike pseudotyped virus showed reduced entry into human Calu-3 lung cells compared to LP.8.1^2^ and low infectivity of Vero cells relative to LP.8.1^11^ . Live BA.3.2 virus was not tested. Therefore, viral properties such as cytopathic effect (CPE) and replication were not measured.

In this work we characterized live BA.3.2 virus which we isolated from a swab from the national South African sentinel surveillance program, as well as its escape from neutralization by blood plasma collected in South Africa at different timepoints during the pandemic. We found that this isolate, along with almost all BA.3.2 sequences to date, had complete deletions of ORF 7 and 8. Phylogenetic analysis revealed a likely Southern African origin for BA.3.2. CPE measured as plaque size was lower relative to Delta, ancestral/D614G SARS-CoV-2, and Omicron BA.1, but similar to LP.8.1. Replication of BA.3.2 was similar to LP.8.1 and higher than ancestral SARS-CoV-2 in H1299-ACE2 cells. BA.3.2 also had moderately higher escape from antibody neutralization in recently collected sera relative to LP.8.1. This data shows that, despite the lower infectivity of BA.3.2 spike reported in pseudovirus assays, BA.3.2 has similar replication properties in cell culture while maintaining a moderate neutralization escape advantage over a co-circulating subvariant evolved from JN.1. It may therefore have the potential to acquire further mutations that may give it a fitness advantage and spread further.

## Results

### Phylogenetic origin of BA.3.2

We examined the phylogenetic origin of BA.3.2. Phylogenetic analysis places BA.3.2 as a direct descendant of BA.3 sequences circulating in Southern Africa in late 2021 to 2022 (Fig 1A), and BA.3.2 viral sequences originating in South Africa are closest to the BA.3.2 root (Fig 1A). The emergence of BA.3.2 is estimated to have happened between December 2023 and July 2024. BA.3.2 therefore harbors substantial diversity that has accumulated over at least one year. As of writing (October 2025), new sequences of BA.3.2 were recently uploaded to GISAID from Australia, indicating persistent circulation (Fig 1A). The reconstructed ancestral BA.3.2 sequence differs from its closest BA.3 ancestor by 36 substitutions, two deletions, and one four amino acid insertion in the spike protein (Fig. 1B).

**Figure 1:**
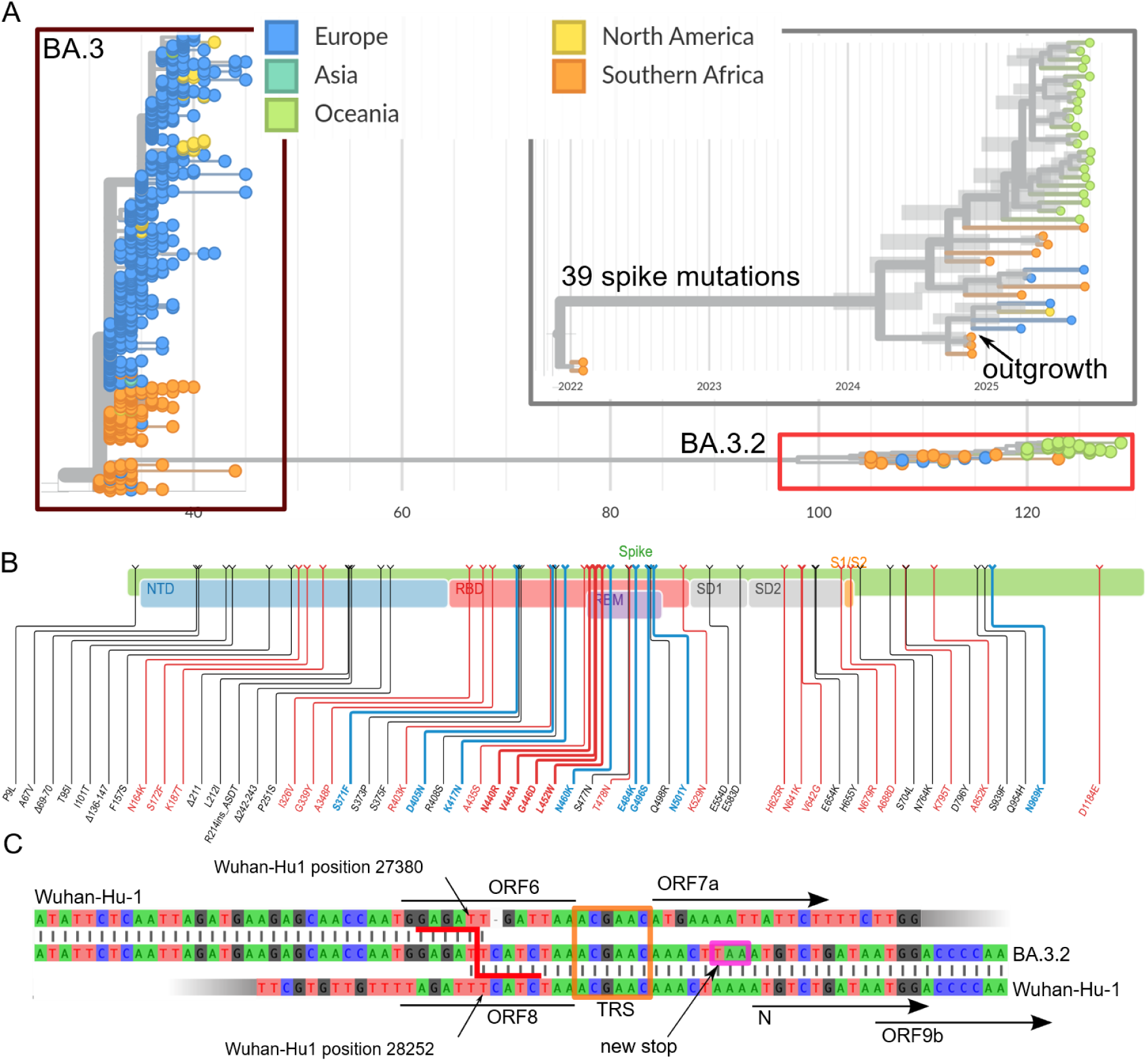
Geographic origin of BA.3.2. A) Phylogenetic tree of sequences colored by geographic origin from BA.3 sequences sampled in late 2021/early 2022 and BA.3.2 sequences from 2024-2025. BA.3.2 is most closely related to BA.3 sequences from Southern Africa and basal BA.3.2 sequences also originated in Southern Africa. Inset: time scaled phylogeny, showing that the common ancestor of BA.3.2 sequences likely circulated in the first half of 2024. B) Summary of amino acid substitutions, insertions, and deletions in Spike of the inferred root of BA.3.2 relative to the Wuhan-Hu1 reference. C) Genetic context of the large deletion removing ORF7a,b and ORF8. The BA.3.2 genome (middle) aligns with the reference until reference position 27380 (top) at the end of ORF6. Then the sequence switches to reference position 28252 at the end of ORF8, resulting in an 871 base deletion comprising ORFs 7a, 7b and 8. ORF6 is terminated by a new stop-codon immediately in front of the start of ORF N (purple box). Deletions in this area are likely facilitated by transcription regulatory sequence (TRS, orange box) identity.

We isolated BA.3.2.1 virus from a nasopharyngeal swab sample collected in Gauteng Province, South Africa in November 2024 from a 5-year-old male child (GISAID accession EPI_ISL_19771107). Interestingly, this genome lacks ORF7 and ORF8 due to an 870-base deletion (Fig 1C). The deletion itself may have been facilitated by sequence similarity between transcription regulatory sites (TRS) upstream of ORF7 and nucleocapsid (N). The deletion also removed the canonical stop codon of ORF6, which in BA.3.2 is terminated by a new stop codon immediately before N (Fig 1C).

This deletion is not reported in the majority (36/40) of genomes shared to date (October 2025). Instead, most genome report a long stretch of the ambiguous character “N” at this location, presumably indicating lack of coverage. Almost all genomes, however, are compatible with the deletion of 873 bases reported in Fig 1C. The lack of reported deletions is likely due to the inability of genome assembly pipelines to correctly process deletions that are much longer than individual reads. We conclude that this deletion is likely present in almost all BA.3.2 sequences to date.

### BA.3.2 cytopathic effect and replication kinetics

To test viral properties, we used H1299 cells overexpressing the ACE2 receptor^12^. H1299 cells have been previously reported to have a functional interferon response^9,13–17^. They have low levels of TMPRSS2 and have been shown to be predominantly infected by SARS-CoV-2 through the endocytic pathway^18^.

We quantified cytopathic effect (CPE) at 72 hours post-infection by plaque size, which represents the area of dead cells resulting from one infectious viral unit. We used a range of low input virus titers, which we determined in the same experiment 24 hours post-infection by a focus forming assay (Fig S2, Methods). Plaque size was quantified by a bespoke image analysis pipeline (Fig 2A, Methods). To compare plaque areas relative to BA.3.2, we calculated the geometric mean of BA.3.2 plaque areas in pixels. We then divided each plaque area for all strains by the BA.3.2 plaque geometric mean. The results show plaque area to be variable within the same viral strain (Fig 2B). However, plaque areas also varied between strains, being largest in the Delta variant infected cells and smallest in JN.1 Omicron subvariant infections (Fig 2B). BA.3.2 had significantly smaller plaques relative to Delta (3.2-fold smaller, p<0.0001, by the Kruskal-Wallis test with the dunn-sidak correction for multiple hypothesis) ancestral virus with the D614G mutation (henceforth D614G, 2.5-fold, p<0.0001), and BA.1 (1.4-fold, p=0.001), but twice larger than JN.1 (p=0.04). BA.3.2 plaque areas were not significantly different relative to those made by LP.8.1 (Fig 2B).

**Figure 2:**
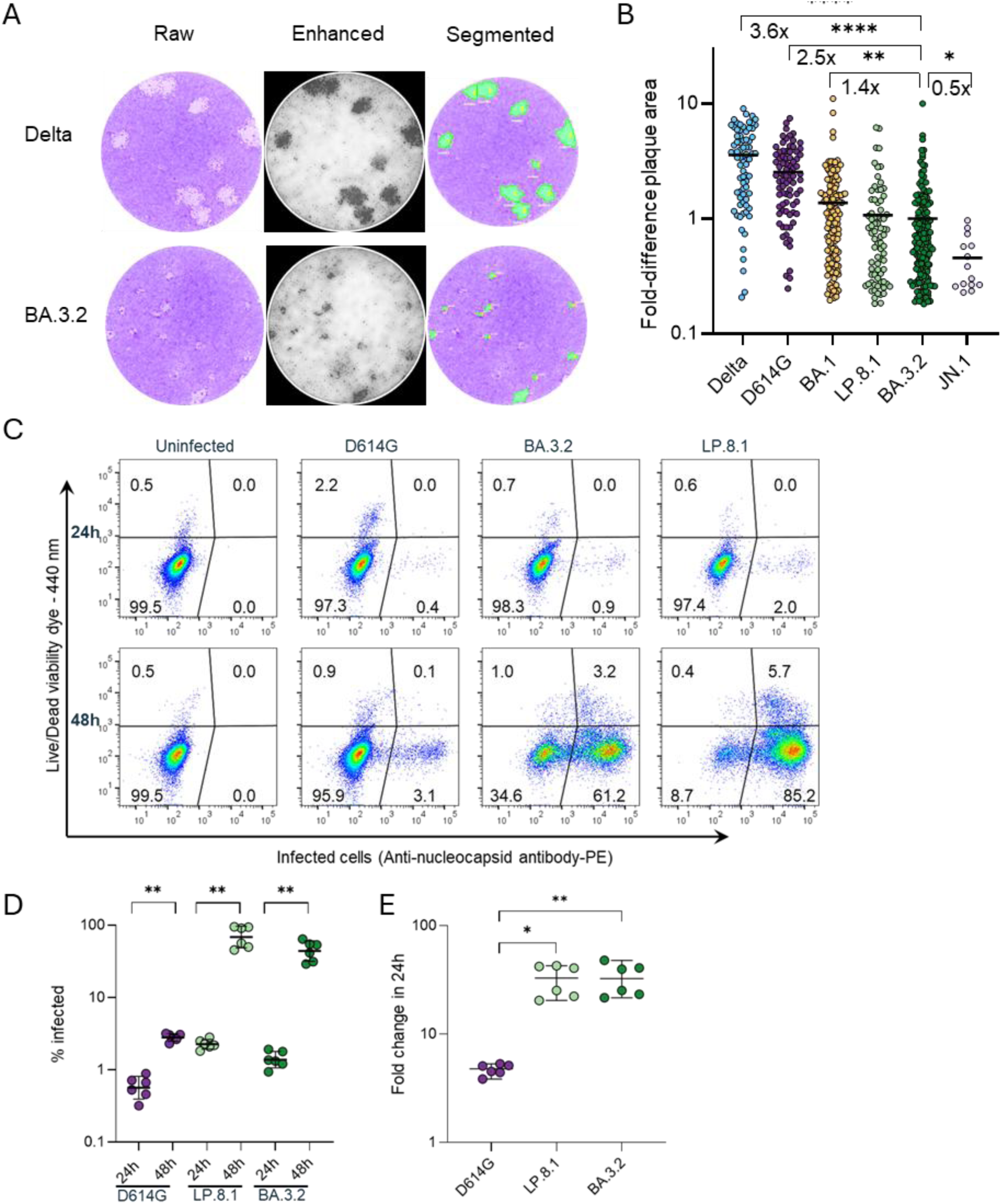
BA.3.2 infection in cell culture. A-B) H1299-ACE2 cells were infected with Delta, D614G, BA.1, BA.3.2, LP.8.1 and JN.1 viruses and plaques quantified 72 hours post-infection. Image analysis pipeline is shown in A and plaque sizes per strain normalized by the geometric mean plaque size for BA.3.2 is shown in B. Bars are geometric means of fold difference versus BA.3.2. Plaque numbers are n=76 for Delta, n=82 for D614G, n=141 for BA.1, n=76 for LP.8.1, n=159 for BA.3.2, n=14 for JN.1 from three independent experiments. *p=0.04, **p=0.001, ****p<10^−4^ determined by the Wilcoxon test with Benjamini–Hochberg multiple hypothesis correction. C) Representative dot plots of flow cytometry of infected cells 24 hours and 48 hours post-infection as detected by anti-SARS-CoV-2 nucleocapsid antibody (x-axis), with y-axis showing live versus dead cells. D) Quantified percentage of infected cells at 24 and 48 hours for D614G, LP.8.1 and BA.3.2 infections. Bars are medians and error bars are 95% confidence intervals from three independent experiments with three repeats each. **p=0.002 by 2-tailed Mann-Whitney test. E) Fold-change in infected cells quantified as percent infected at 48 hours divided by median infection at 24 hours. *p=0.01, **p=0.009 by the two-tailed Kruskal–Wallis test with Dunn multiple hypothesis correction.

We also measured viral replication in H1299-ACE2 cells over 24 hours. For this, we compared the proportion of infected cells at 24 versus 48 hours after the same input infection dose to quantify the fold-change in infection over a 24-hour time period. We detected SARS-CoV-2 positive cells at 24 and 48 hours using flow cytometry by an antibody to the SARS-CoV-2 nucleoprotein (Fig 2C, Methods). We tested ancestral SARS-CoV-2 with D614G, LP.8.1, and BA.3.2. At 24 hours, H1299-ACE2 cells infected with D614G showed a median infection of 0.6%, increasing to a median of 2.9% at 48h (significant increase, p=0.002 by the Mann-Whitney non-parametric test, Fig 2D), while LP.8.1 increased from a median of 2.2% to 74% (p=0.002) and BA.3.2 increased from a median of 1.3% to 48% (p=0.002). To calculate fold-change, we divided the percent infected cells after 48h infection by the median percentage at 24h per strain. Both BA.3.2 and LP.8.1 had a median 33-fold increase over 24 hours (Fig 2E). This was significantly higher replication relative to D614G, which showed a median 4.8 fold-increase over the 24h time period (p=0.009 for BA.3.2, p=0.01 for LP.8.1 by the Kruskal-Wallis test with the dunn-sidak correction for multiple hypothesis). Hence, BA.3.2 and LP.8.1 had similar replication in this cell line which was faster than D614G.

### Immune escape through time

To test BA.3.2 escape from neutralizing antibodies and compare it to other SARS-CoV-2 subvariants, we used plasma from participants collected at different times in the pandemic in South Africa (Table 1). The first set of plasma samples was collected before Omicron arose at the end of 2021. This plasma was from convalescent participants about 1-month post-infection, where infection was by D614G, Beta, or the Delta variant of SARS-CoV-2^19–21^. No participant in this group was vaccinated before blood was collected.

**Table 1:**
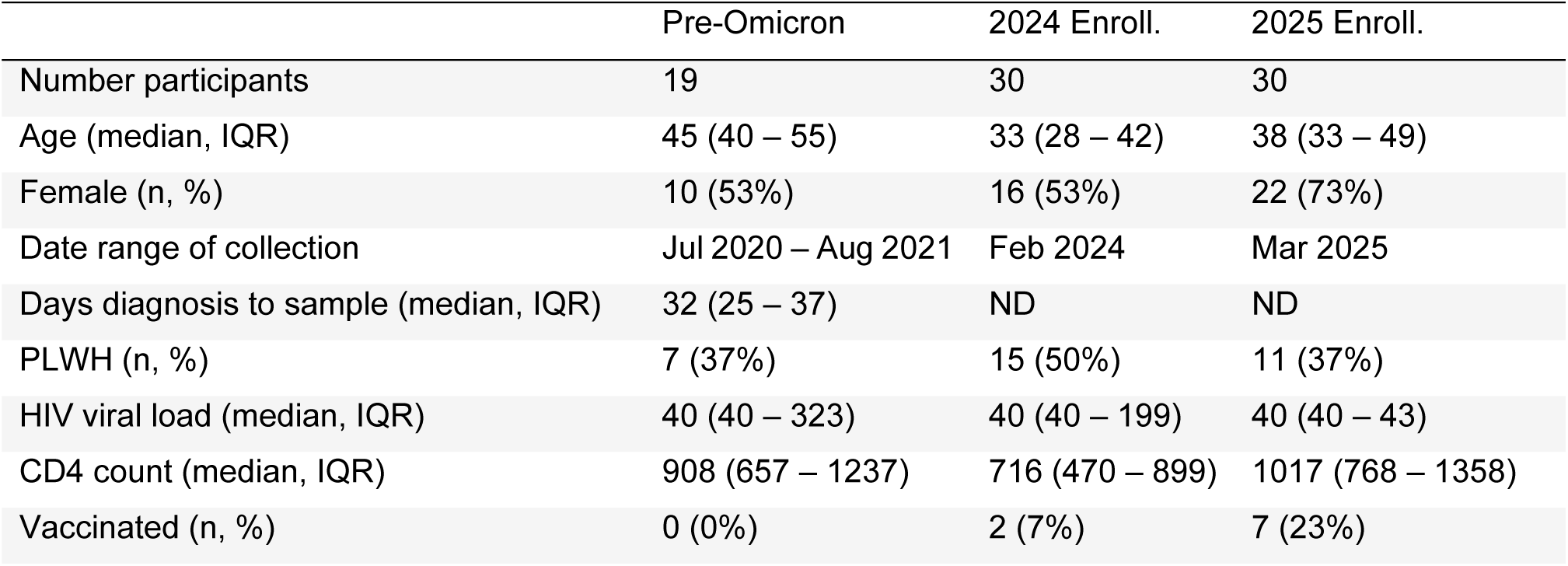
Participant characteristics of plasma used in neutralization experiments.

|  | Pre-Omicron | 2024 Enroll. | 2025 Enroll. |
| --- | --- | --- | --- |
| Number participants | 19 | 30 | 30 |
| Age (median, IQR) | 45 (40 – 55) | 33 (28 – 42) | 38 (33 – 49) |
| Female (n, %) | 10 (53%) | 16 (53%) | 22 (73%) |
| Date range of collection | Jul 2020 – Aug 2021 | Feb 2024 | Mar 2025 |
| Days diagnosis to sample (median, IQR) | 32 (25 – 37) | ND | ND |
| PLWH (n, %) | 7 (37%) | 15 (50%) | 11 (37%) |
| HIV viral load (median, IQR) | 40 (40 – 323) | 40 (40 – 199) | 40 (40 – 43) |
| CD4 count (median, IQR) | 908 (657 – 1237) | 716 (470 – 899) | 1017 (768 – 1358) |
| Vaccinated (n, %) | 0 (0%) | 2 (7%) | 7 (23%) |

More recently, it has been difficult to enroll confirmed SARS-CoV-2 cases since testing has been discontinued. Therefore, to assess more recent antibody immunity, enrolled participants without SARS-CoV-2 symptoms and without knowing their infection history. Even pre-Omicron, seroprevalence of SARS-CoV-2 infection in South Africa was about 60-70%^22^ and we therefore expected that the more recently enrolled participants would have an exposure to SARS-CoV-2 through infection, vaccination, or both. Enrollment for the first group was in February 2024, when JN.1 was circulating. Enrollment for the second group was in March 2025, when LP.8.1 was circulating (Fig 3A). A minority were vaccinated (Table 1). HIV prevalence, which was between 37 to 50%, was consistent with the previously reported high prevalence in the KwaZulu-Natal area where our study is based^23,24^, although measured median CD4 counts and HIV viral loads indicated that participants living with HIV had controlled HIV infections (Table 1).

**Figure 3:**
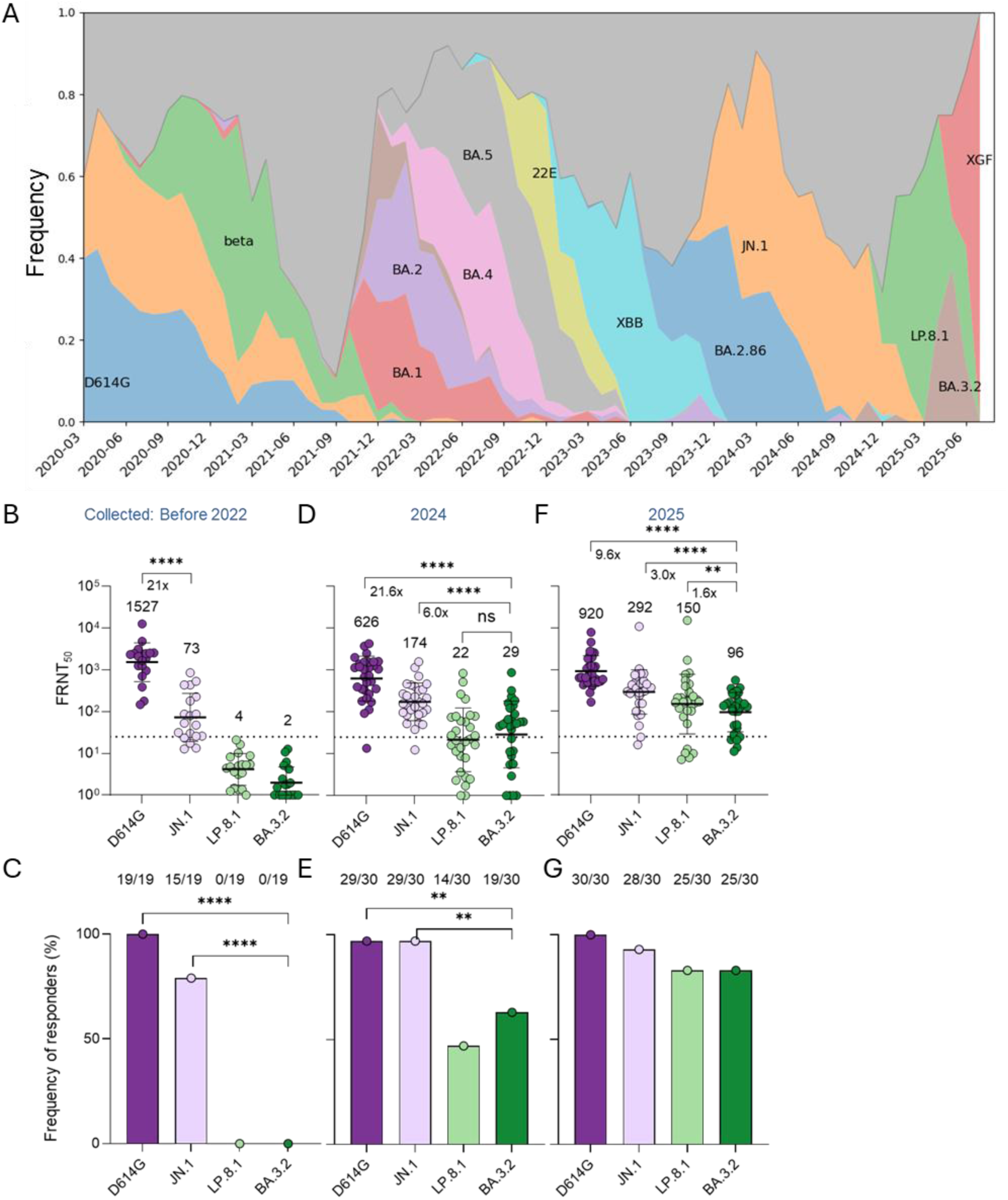
BA.3.2 neutralization at different times in the pandemic. A) Variant frequency in South Africa over time. B) D614G, JN.1, LP.8.1 and BA.3.2 neutralization by pre-2022 plasma from infected unvaccinated South Africans. Bars are geometric mean titers. ****p<10^−4^ by 2-tailed Wilcoxon test. C) Frequency of responders in B, defined as participants able to neutralize above the limit of quantification (FRNT_50_=25, the most concentrated plasma used). ****p<10^−4^ by Fisher’s Exact test. D) Neutralization by February 2024 collected plasma from South Africans with no Covid-19 symptoms. ****p<10^−4^ by 2-tailed Wilcoxon test. E) Frequency of responders in D. **p=0.003 by the Fisher’s Exact test. F) Neutralization by March 2025 collected plasma from South Africans with no Covid-19 symptoms. ****p<10^−4^, **p=0.006 by 2-tailed Wilcoxon test. G) Frequency of responders in F.

To quantify neutralization, we used the focus reduction neutralization 50% titer (FRNT_50_), the inverse of the plasma dilution which neutralized 50% of the virus as detected in a live virus focus forming assay. Plasma collected pre-Omicron showed strong neutralization of D614G virus (Fig 3B) with all participants able to neutralize this strain (Fig 3C) above the limit of quantification (FRNT_50_=25, corresponding to a 1:25 plasma dilution, the most concentrated plasma used). JN.1 neutralization was 21-fold lower for these participants. However, as previously reported for the BA.1 subvariant^19^, escape was not complete, with 15 out of 19 tested participants (79%) able to neutralize the virus above the limit of quantification (Fig 3C). Neutralization of LP.8.1 and BA.3.2 was below level of quantification for all participants (Fig 3C).

Plasma samples collected from 2024 participants showed higher neutralization of JN.1 and lower neutralization of D614G relative to plasma of pre-Omicron participants (Fig 3D). The fraction of neutralizers increased for JN.1 to 29 out of 30 (97%) and was the same as for D614G (Fig 3E). neutralization of LP.8.1 and BA.3.2 increased to low but detectable levels (Fig 3D), with 19/30 participants showing an ability to detectably neutralize BA.3.2 (Fig 3E). This proportion was not significantly different from LP.8.1 (Fig 3E). BA.3.2 showed 21.6-fold lower neutralization relative to D614G and 6-fold lower neutralization relative to JN.1 (Fig 3D).

In 2025, there was a further increase in neutralization capacity against both LP.8.1 and BA.3.2 (Fig 3F), and 25/30 of the plasma samples neutralized BA.3.2 above the limit of quantification (Fig 3G). However, neutralization of this subvariant was significantly lower than D614G (by 9.6-fold) and JN.1 (3-fold). It was also moderately but significantly lower than LP.8.1 (1.6-fold), with LP.8.1 circulating at the time of collection (Fig 3A).

Antigenic cartography of the viral strains against all plasma samples showed D614G virus to be at the center, surrounded by plasma samples from the three participant groups – it was the most neutralized virus across time. The virus with the shortest antigenic distance from D614G was JN.1, while BA.3.2 and LP.8.1 were antigenically removed from both D614G and JN.1 (Fig 4). With time, plasma samples reduce antigenic distance to both LP.8.1 and BA.3.2.

**Figure 4:**
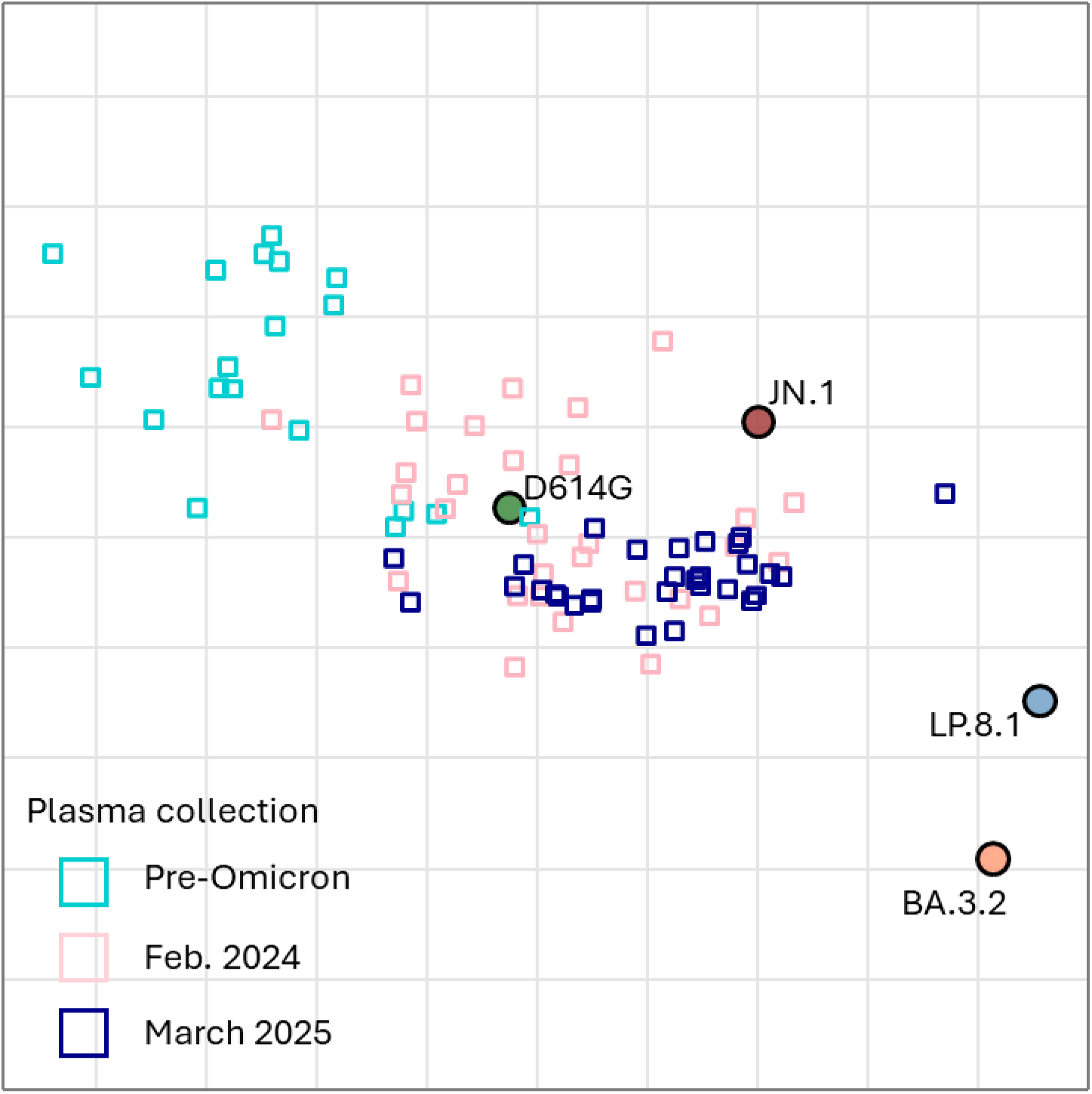
Antigenic cartography shows separation of BA.3.2 from plasma collected at different times in the pandemic. Circles are viruses, small squares are plasma samples, and each square on the grid is a 2-fold drop in neutralization. Pre-omicron plasma samples are shown in cyan, 2024 samples in pink, and 2025 samples in dark blue.

## Discussion

In this study we investigated the geographic origin and properties of the Omicron BA.3.2 subvariant, which was isolated as the BA.3.2.1 sub-lineage. The putative ancestor BA.3.2 has not been sampled. It is unclear whether BA.3.2.1 and BA.3.2.2 emerged from one infected individual, or whether this diversity is due to low level circulation in the population. We found that BA.3.2 likely evolved in Southern Africa in the first half of 2024. The Southern African region gave rise to other variants and subvariants like Beta, BA.1, BA.2, BA.3, and BA.2.86, the precursor to JN.1. Beta showed extensive infection in the Southern African region, with high morbidity and mortality^12,23^, while BA.1, BA.2, and JN.1 spread worldwide. One explanation for the region being a hotspot for SARS-CoV-2 variant evolution is the high prevalence of HIV infection. Since about 10% of people living with HIV may not effectively suppress the virus with antiretroviral drugs because of lack of access or adherence to antiretroviral therapy, the high HIV prevalence translates to a large group of people with immunocompromise due to advanced HIV disease^1,25^. Our previous work and that of others showed persistent SARS-CoV-2 infections and extensive SARS-CoV-2 evolution in immunocompromised people, including those with advanced HIV disease^20,26–35^.

For neutralization, our results were broadly consistent with previous studies^2,11^ showing BA.3.2 to have strong escape from neutralizing antibodies elicited by previous SARS-CoV-2 exposures. However, we also observed that neutralizing antibody immunity against BA.3.2 increased with time, and that most 2025 plasma samples from participants enrolled in the geographical region in which BA.3.2 likely evolved show detectable neutralization of this subvariant. However, neutralization of BA.3.2 is still significantly lower than the co-circulating LP.8.1 subvariant. One possible explanation is that, since LP.8.1 was circulating at relatively high levels in South Africa relative to BA.3.2, some of our participants may have been infected with LP.8.1 and developed specific immunity to it.

We found that almost all reported BA.3.2 sequences are compatible with the deletion of 873 bases which results in the deletion of ORF8, as well as ORF7a and ORF7b. ORF8 is reported to interfere in type 1 interferon signalling^36^ as well as downregulate MHC-I^37^. ORF8 also increases ER stress which may accelerate cell death^38^. Deletion of ORF8 is common^39^ and may be positively selected in SARS-CoV-2 Omicron subvariants^40^. In one study, individuals infected with SARS-CoV-2 with an ORF8 deletion mutant were reported to show milder disease^41^, although other studies showed little effect of an ORF8 deletion on replicative fitness^42^ or host transcriptional response^43^. It is tempting to speculate that these deletions attenuate the virus, but disease severity is complex and depends not only on the virus genotype but on the level of SARS-CoV-2 immunity in the host^44,45^ as well as host risk factors such as co-morbidities^46^. More epidemiological data is required.

BA.3.2 showed similar cytotoxicity and replication to LP.8.1 in H1299-ACE2 cells despite the previously reported reduced entry^2^ and low infectivity^11^ relative to LP.8.1. Delta and ancestral SARS-CoV-2, which have been reported to cause more severe disease relative to Omicron subvariants^44^, show cell culture phenotypes such as increased cytotoxicity relative to Omicron^9,47,48^. This is consistent with our results. On the other hand, Omicron subvariants infect better than ancestral SARS-CoV-2 through the endocytic pathway^18,47^ and would be expected to show faster replication relative to ancestral SARS-CoV-2 in H1299 cells predominantly infected through the endocytic pathway^18,49,50^, as we observed in this study. With the caveat that the infection phenotype in cell culture is a very rough indication of what the virus can do on the organism level, the similarity we observed in terms of cytotoxic effect and replication between BA.3.2 and LP.8.1 does not give an indication that BA.3.2 is potentially more pathogenic than LP.8.1.

Limitations of the study include the undefined infection history of participants enrolled in 2024 and 2025. In addition, H1299-ACE2 cells, while convenient for assays such as CPE since they form well defined and reproducible monolayers^19^, may not reflect infection in cells with endogenous ACE2 levels, primary cells with intact regulatory pathways, or cells where the main infection route is thorough the plasma membrane by the TMPRSS-2 dependent spike cleavage as opposed to the endocytic pathway^18,50^.

BA.3.2 infections have been expanding slowly, and the potential of BA.3.2 or its descendants to further evolve and expand is unclear. The BA.2.86 variant showed slow expansion until it evolved the L455S mutation to become JN.1, which spread globally. More broadly, the continual evolution of long branch variants in Southern Africa may point to continuing SARS-CoV-2 evolution in persistent infections in people immunocompromised because of advanced HIV disease^1^. Erosion of the ability to deliver antiretroviral therapy to people living with HIV may increase the number of people with advanced HIV disease and lead to further evolution of SARS-CoV-2 and other viruses.

## Methods

### Informed consent and ethical statement

All blood samples used for neutralization studies, as well as nasopharyngeal swabs for isolation of the ancestral/D614G and Delta virus were collected from adults (>18 years old) enrolled at King Edward VIII, Inkosi Albert Luthuli Central, or Clairwood hospitals or the University of KwaZulu-Natal Medical School Campus in Durban, South Africa. Written informed consent was obtained. Study was approved by the Biomedical Research Ethics Committee at the University of KwaZulu-Natal (reference BREC/00001275/2020). The Omicron BA.1, JN.1, LP8.1 and BA.3.2 viruses were isolated from residual swabs used for diagnostic testing, as approved by the Witwatersrand Human Research Ethics Committee (reference M210752).

### Data and reagent availability statement

Viral isolates are available upon reasonable request to the authors. Sequences of isolated SARS-CoV-2 used in this study have been deposited in GISAID with accession numbers as follows:

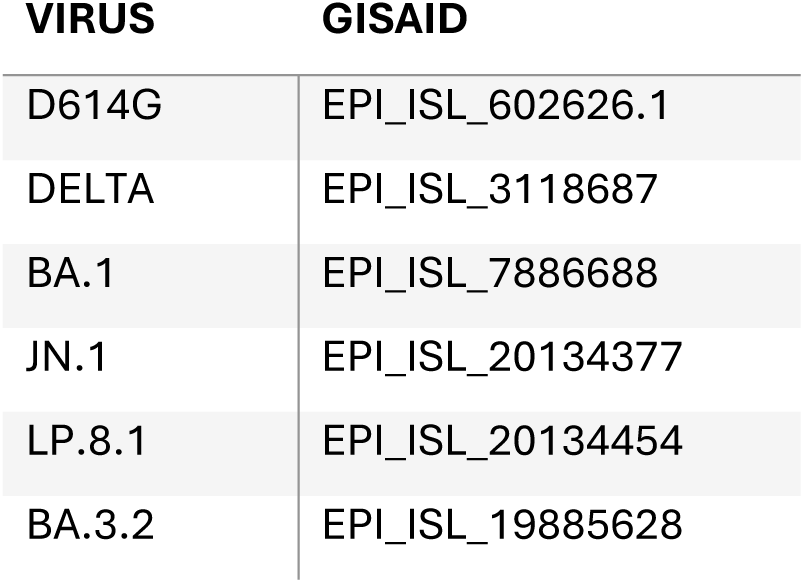

All data produced in the present study are available upon reasonable request to the authors.

### Clinical laboratory testing

HIV viral load quantification was performed at an accredited diagnostic laboratory (Molecular Diagnostic Services, Durban, South Africa). The CD4 count was performed by an accredited diagnostic laboratory (Ampath, Durban, South Africa).

### Whole-genome sequencing

Sequencing was performed on an Illumina MiSeq or NextSeq 550 instrument, or alternatively the Oxford Nanopore instrument. For Illumina sequencing, total nucleic acid was extracted on an automated Chemagic 360 instrument, using the CMG-1033-S kit (Perkin Elmer, Hamburg, Germany). Libraries for whole genome sequencing were prepared using the Illumina COVIDseq Assay as per manufactures instructions. For the Illumina COVIDseq assay, amplicons were tagmented, followed by indexing using the Illumina UD Indexes. Sequencing libraries were pooled and normalized to 4 nM. An 8 pM sample library was spiked with 1% PhiX (PhiX Control v3 adaptor-ligated library used as a control). Libraries were sequenced using a 500-cycle v2 MiSeq Reagent Kit on the Illumina MiSeq instrument (Illumina Inc, USA). On the Illumina NextSeq 550 instrument, sequencing was performed using the Illumina COVIDSeq protocol (Illumina Inc, USA). First strand synthesis was done using random reverse transcriptase and hexamer primers. The SARS-CoV-2 genome was amplified using the Artic V5.4.2 NCOV-2019 Panel, Pool 1 and 2. The pooled PCR amplified products were processed for tagmentation and adapter ligation done with Illumina UD indexes. Further enrichment and cleanup was performed as per manufacturer’s instructions (Illumina Inc, USA). Pooled samples were quantified using Qubit 4.0 fluorometer (ThermoFisher Scientific, Oregon, USA) using the Qubit dsDNA High Sensitivity assay according to manufacturer’s instructions. The fragment sizes were analyzed using TapeStation 4200 (Agilent Technologies, Santa Clara, CA). Pooled libraries were normalized to 4nM concentration and 25 μL of each normalized pool containing unique index adapter sets were combined in a new tube. The final library pool was normalized to 0.65 pM and spiked with 10% PhiX. Libraries were loaded onto a 300-cycle P1 NextSeq reagent kit (Illumina Inc, USA) and ran on the Illumina NextSeq 1000/2000 instrument (Illumina, San Diego, CA, USA). For Illumina assembly, GATK HaploTypeCaller --min-pruning 0 argument was added to increase mutation calling sensitivity near sequencing gaps. For Oxford Nanopore sequencing, the Midnight Primer kit (Oxford Nanopore Technologies, UK) was used as described by Freed and Silander (2020)^51^. RNA extraction was done using the QIAamp Viral RNA Mini kit as per manufacturer’s instructions (Qiagen, Germany). cDNA synthesis was performed on the extracted RNA using LunaScript RT mastermix (New England Biolabs, USA) followed by gene-specific multiplex PCR using the Midnight Primer pools which produce 1200bp amplicons which overlap to cover the 30-kb SARS-CoV-2 genome. Amplicons from each pool were pooled and used neat for barcoding with the Oxford Nanopore Rapid Barcoding kit (Oxford Nanopore Technologies, UK) as per the manufacturer’s protocol. Barcoded samples were pooled and bead-purified. After the bead clean-up, the library was loaded on a prepared R10.4.1 flow-cell. A MinION sequencing run was initiated using MinKNOW software with the base-call setting switched off. We assembled paired-end and nanopore.fastq reads using Genome Detective 2.21.3 (https://www.genomedetective.com) which was updated for the accurate assembly and variant calling of tiled primer amplicon Oxford Nanopore reads, and the Coronavirus Typing Tool. Additionally, the ARTIC SARS-CoV-2 pipeline (wf-artic) from EPI2ME Labs was executed using the Nextflow workflow framework. The reference genome used throughout the assembly process was NC_045512.2 (numbering equivalent to MN908947.3). To determine clade assignment, mutation changes and quality scores, consensus FASTA files were entered into Nextclade (https://clades.nextstrain.org/). Mutations were then visualized in Microsoft Excel relative to the infecting variant.

### Phylogenetic analysis

Phylogenetic analysis is based on EPI_SET_251022ev. We assembled a set of 727 BA.3 (Nextstrain clade 21M) sequences collected between November 2021 and June 2022 from data deposited on GISAID^52^. BA.3.2 sequences were downloaded on 2025-10-15 directly from GISAID. We excluded sequences flagged as poor quality by Nextclade^53^. Sequences were pairwise aligned against Wuhan-Hu-1 using Nextclade. A tree was built using IQ-tree 2^54^ and postprocessed using a custom script to correct for incomplete merging of branches in large polytomies. A time tree was inferred by TreeTime^55^ using a clock rate of 0.0005 per site and year^56^. The rate of the long branch between BA.3 and BA.3.2 was set to be 2 times the rate of the rest of the tree in line with previous observation that evolution is 2-fold accelerated along many long branches leading to distinct clades. This acceleration is consistent with the strong enrichment of amino acid substitutions in the spike protein along the long branch leading to BA.3.2, suggesting that evolution along this branch is dominated by adaptive changes. The full workflow to reproduce the analysis is available at https://www.github.com/neherlab/BA_3_2. The repository contains a specific list of sites (config/mask.tsv) that are masked in individual sequences due to suspicion of being artefactual reversions.

### Cells

The VeroE6 cells expressing TMPRSS2 and ACE2 (VeroE6-TMPRSS2), originally BEI Resources, NR-54970 were used for virus expansion and all live virus assays excluding replication and plaque assay. The Vero-TMPRSS2 cell line was propagated in growth medium consisting of Dulbecco’s Modified Eagle Medium (DMEM, Gibco 41965-039) with 10% fetal bovine serum (Hyclone, SV30160.03) containing 10mM of hydroxyethylpiperazine ethanesulfonic acid (HEPES, Lonza, 17-737E), 1mM sodium pyruvate (Gibco, 11360-039), 2mM L-glutamine (Lonza BE17-605E) and 0.1mM nonessential amino acids (Lonza 13-114E). The H1299-E3 (H1299-ACE2, clone E3) cell line used in the plaque and replication assays was derived from H1299 (CRL-5803) and propagated in growth medium consisting of complete Roswell Park Memorial Institute (RPMI, Gibco, 21875-034) 1640 with 10% fetal bovine serum containing 10mM of HEPES, 1mM sodium pyruvate, 2mM L-glutamine and 0.1mM nonessential amino acids.

### Virus expansion

All work with live virus was performed in Biosafety Level 3 containment using protocols for SARS-CoV-2 approved by the Africa Health Research Institute Biosafety Committee. VeroE6-TMPRSS2 cells were seeded at 4.5 × 10^5^ cells in a 6 well plate well and incubated for 18–20 hours pre-infection. After one Dulbecco’s phosphate-buffered saline (DPBS) wash, the sub-confluent cell monolayer was inoculated with 500μL of universal transport medium which contained the swab, diluted 1:2 with growth medium filtered through a 0.45μm filter. Cells were incubated for 2 hours at 37 °C, 5% CO2. Wells were then filled with 3 mL complete growth medium. After 3 days of infection (completion of passage 1 (P1)), supernatant was collected, cells were trypsinized, centrifuged at 300 × *g* for 3 min and resuspended in 3mL growth medium. All infected cells and supernatant were added for cell-to-cell infection to VeroE6-TMPRSS2 cells, with medium removed, that had been seeded at 1.5 × 10^5^ cells per mL, 3 × 10^6^ cells total, 18–20 hour earlier in a T75 flask. The coculture was incubated for 2 h and the flask was filled with 20mL of complete growth medium and incubated for 3 days. The viral supernatant from this culture (passage 2 (P2) stock) was used for experiments.

### Plaque assay

H1299-E3 cells were plated in a 96-well plate (Corning) at 20,000 cells per well 1 day pre-infection. Virus stocks (used at the 50 focus-forming units per microwell and serially diluted 1:2) were added to cells, incubated for 1 h at 37 °C, 5% CO_2_. Following incubation, 100μL of a 1X RPMI 1640 (Sigma-Aldrich, R6504), 1.5% carboxymethylcellulose (Sigma-Aldrich, C4888) overlay was added without removing the inoculum. Cells were fixed 72 hours post-infection using 4% PFA (Sigma-Aldrich, P6148) for 20 min. The fixed cells were washed with buffer containing 0.05% Tween-20 in PBS and stained with 30 μL/well of a 0.5% crystal violet solution (Sigma-Aldrich, 61135). After staining, cells were washed with distilled water. Plates were imaged in an ImmunoSpot Ultra-V S6-02-6140 Analyzer ELISPOT instrument with BioSpot Professional built-in image analysis (C.T.L).

### Image analysis to determine individual plaque area

Plaque sizes were quantified from well images using Matlab R2025a. First, images were converted from RGB to HSV and the saturation channel was used to convert this image to grayscale. Greyscale images were then preprocessed by stretching along the interval [0.01 0.99] then adjusted using the Matlab imadjust function with gamma=2. A mask of this image was created and the image was blurred using an averaging filter in the masked (bright) image region. A second mask was generated of the filtered image with pixels in well perimeter assigned “1” values. This mask was inverted, showing pixels in non-plaque regions and well perimeter as “0” values. This mask was used to segment the plaques, with watershed segmentation separating overlapping plaques. For improved accuracy, manual curation was performed by redrawing individual plaques where automatic segmentation was insufficient or to exclude pipette scratches or other artifacts.

### Replication assay by flow cytometry

H1299-E3 cells were plated at 1 × 10^5^cells per well in 12-well plates (Corning) 1-day pre-infection. The following day, cells were infected with 50 focus-forming units in 200 μL growth media per well. Cell-virus mixtures were incubated for 1 h at 37 °C, 5% CO2 followed by 1 mL of growth media. The following days, 24- and 48-hours post-infection, cells were trypsinized, collected and stained with Blue Live/Dead stain as per manufacturer instructions (L34961, ThermoScientific). The samples were then washed in 1 mL PBS and resuspended in Cytofix/Cytoperm buffer (BD Biosciences) for 20 min at 4⁰C in the dark. The samples were then stained with 0.5 μg/mL anti-SARS-CoV-2 nucleocapsid-PE (ab283244, Abcam) diluted with Perm/Wash buffer (BD Biosciences) containing Fetal Bovine Serum (FBS) and saponin for 1 hour at 4⁰C in the dark. Cells were analysed on an BD FACSymphony. Data was analysed using FlowJo V10.10.0 and Graphpad Prism V10 software

### Live virus focus forming assay and neutralization assay

For all neutralization assays, viral input was 100 focus forming units per well of a 96-well plate. VeroE6-TMPRSS2 cells were plated in a 96-well plate (Corning) at 20,000 cells per well 1 day pre-infection. Plasma was separated from EDTA-anticoagulated blood by centrifugation at 500 × *g* for 10 min and stored at −80 °C. Aliquots of plasma samples were heat-inactivated at 56 °C for 30 min and clarified by centrifugation at 10,000 × *g* for 5 min. Virus stocks were added to diluted plasma in neutralization assays. Antibody–virus mixtures were incubated for 1 h at 37 °C, 5% CO_2_. Cells were infected with 100 μL of the virus–antibody mixtures for 1 h, then 100 μL of a 1X RPMI 1640 (Sigma-Aldrich, R6504), 1.5% carboxymethylcellulose (Sigma-Aldrich, C4888) overlay was added without removing the inoculum. Cells were fixed 20 h post-infection using 4% PFA (Sigma-Aldrich, P6148) for 20 min. Foci were stained with a rabbit anti-spike monoclonal antibody (BS-R2B12, GenScript A02058) at 0.5 μg/mL in a permeabilization buffer containing 0.1% saponin (Sigma-Aldrich, S7900), 0.1% BSA (Biowest, P6154) and 0.05% Tween-20 (Sigma-Aldrich, P9416) in PBS for 2 h at room temperature with shaking, then washed with wash buffer containing 0.05% Tween-20 in PBS. Secondary goat anti-rabbit HRP conjugated antibody (Abcam ab205718) was added at 1 μg/mL and incubated for 2 h at room temperature with shaking. TrueBlue peroxidase substrate (SeraCare 5510-0030) was then added at 50 μL per well and incubated for 20 min at room temperature. Plates were imaged in an ImmunoSpot Ultra-V S6-02-6140 Analyzer ELISPOT instrument with BioSpot Professional built-in image analysis (C.T.L) which was also used to quantify areas of individual foci.

### Statistics and fitting

All statistics performed in GraphPad Prism version 9.4.1. All fitting to determine FRNT_50_ and linear regression was performed using custom code in MATLAB R2025a (FRNT_50_) or the fitlm function for linear regression, which was also used to determine goodness-of-fit (R^2^) as well as p-value by F-test of the linear model.

Neutralization data were fit to:

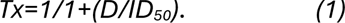

Here Tx is the number of foci at plasma dilution D normalized to the number of foci in the absence of plasma on the same plate. ID_50_ is the plasma dilution giving 50% neutralization.

FRNT_50_ = 1/ID_50_. Values of FRNT_50_ <1 are set to 1 (undiluted), the lowest measurable value. We note that the most concentrated plasma dilution was 1:25 and therefore FRNT_50_ < 25 were extrapolated.

## Data Availability

Viral isolates are available upon reasonable request to the authors. Sequences of isolated SARS-CoV-2 used in this study have been deposited in GISAID with accession numbers as follows:
VIRUS GISAID
D614G EPI_ISL_602626.1
DELTA EPI_ISL_3118687
BA.1 EPI_ISL_7886688
JN.1 EPI_ISL_20134377
LP.8.1 EPI_ISL_20134454
BA.3.2 EPI_ISL_19885628
All data produced in the present study are available upon reasonable request to the authors.

## Acknowledgements

This study was supported by the Wellcome Trust Award 226137/Z/22/Z (AS). The funder had no role in study design, data collection and analysis, decision to publish, or preparation of the manuscript. We gratefully acknowledge originating and submitting laboratories of the genetic sequence and metadata made available through INSDC or GISAID on which the phylogenetic analysis in this paper is based (EPI_SET_251022ev). We also gratefully acknowledge the public and private laboratories in South Africa sharing SARS-CoV-2 specimens for sequencing as part of national genomic for SARS-CoV-2 which enabled both the phylogenetic analysis and the isolation of live SARS-CoV-2 strains from the residual samples.

---

**Figure S1:**
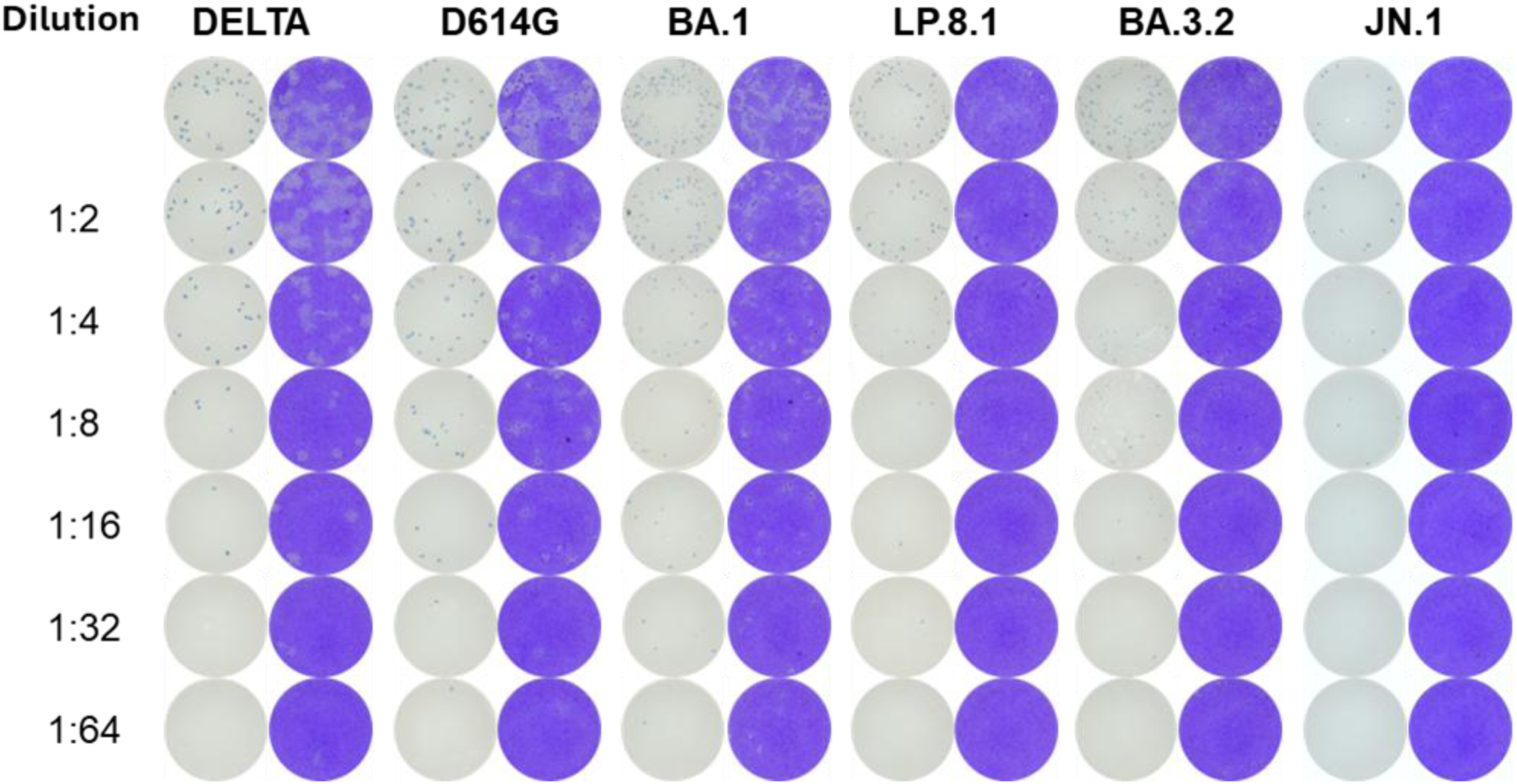
Unprocessed images of plaques and input virus detected by focus forming assay. A 2-fold dilution of input virus was performed for Delta, D614G, BA.1, LP.8.1, BA.3.2, and JN.1 and visualized at 24 hours using a focus forming assay (left column for each virus). Plaques were detected at 72 hours for each dilution (right column for each virus) and dilutions with low numbers of overlapping plaques were used to quantify plaque areas.

